# Predicting community-acquired pneumonia outcome using time series data and machine learning

**DOI:** 10.1101/2025.03.11.25323764

**Authors:** Daniel Lozano-Rojas, Matthew Richardson, Gerrit Woltmann, Robert C. Free

## Abstract

**Background:** Community-acquired pneumonia (CAP) is an acute respiratory condition associated with high mortality in adult populations and is potentially more serious in older patients. Accurate and consistently applied prediction of outcome may contribute to reduce in-hospital mortality. Currently, CAP outcomes are assessed with clinical scores like CURB65, based on signs and symptoms that are non-specific to the disease. Recent literature has shown that machine learning (ML) has the potential to improve outcome prediction, but the sparse and incomplete nature of the data present a challenge for the development of models that can be implemented clinically.

**Methods:** This study aimed to developed ML models that can support outcome prediction in hospital admissions with CAP using routinely collected and time-dependent data from Leicester hospitals. Thus, by modelling mortality prediction, and predicting URB65 on the third day of admission with the forecast of vital signs, implementing a methodology that explores how different characteristics involved in the training process influence the results of the predictions.

**Results:** Data comprised 9390 admissions in the training set, and 7892 in the validation set, for thirty-four clinical variables (fifteen time-dependent). Results of CAP mortality modelling reported AUC of 0.77 using a GRU model that was trained with the time series of vital signs and blood test. Results also showed improvement in models when balancing classes of the target variable in the training set, as well as improvement when using time dependent data. And importantly when predicting URB65 accuracy of 0.85 was obtained when modelled using GRU, when time series were processed using local scaling.

**Conclusions:** This approach might represent an opportunity to anticipate adverse outcomes. These results suggest that ML models utilising time series can have sizable impact in the prediction of CAP outcome, from many perspectives: Complementing currently applied scoring systems approaches like CURB65 in hospital settings, prediction of mortality or forecasting the severity of patients from vital signs that have shown correlation with CAP mortality. The models presented require further validation and development, although they present important indication for CAP mortality prediction.

## Background

Pneumonia is an acute inflammatory condition of the lungs, usually caused by bacterial or viral infection which is among the top ten leading causes of mortality worldwide [1]. It is a substantial public health challenge, contributing not only to high mortality rates but also straining hospital admission resources and Intensive Care Unit (ICU) capacity. Community acquired pneumonia (CAP) results in long-term complications for patients, and imposing substantial financial burdens on healthcare systems [3–6]. In the UK, from 2023-24 there were over 260,000 admissions due to pneumonia, and the NHS reported £1 billion costs associated with pneumonia [7–8].

CAP is a type of pneumonia that affects individuals aged 16 and above who contract an infection before hospital admission. CAP is a particular challenge for older people and makes up most pneumonia cases in the UK [6, 8]. Early treatment of CAP is crucial as delayed treatment can lead to adverse outcomes, although these can be improved with proper management and early detection [5]. In the UK, guidelines support diagnostic confirmation of CAP using chest X-rays. Severity is then assessed using a risk score (CURB65) based on the patient’s symptoms, physiological signs and age. The severity score dictates the level of antibiotics administered and inform the decision where treatment takes place (e.g., intermediate or intensive care units) [9–11]. While the CURB65 score predicts overall mortality, it can underestimate severity in younger patients with low or moderate disease score. Furthermore, it is based on non-specific symptoms and observed values at presentation to admissions, and not on the progression of the infection or response to intervention [12]. Thus, there is a clear need for novel approaches which can provide more nuanced predictive tools, such as machine learning (ML) [13].

ML models are mathematical expressions that use data to identify patterns in structured, and unstructured datasets [14]. These models are then trained using existing data to classify new data using these identified patterns [15]. Such techniques have gained popularity due to their performance for many problems across different fields, such as image recognition, anomaly detection, mortality prediction, and others [16–19].

In our recent review, we showed that while use of ML has been explored in CAP, this has focussed on diagnosis [13] and there has been less research into ML techniques for predicting severity and progression; particularly the use of electronic health records to predict outcomes such as mortality, ICU admission or length of stay (LOS). Additionally, multiple challenges remain, including availability of data for replicability, unbalanced data sets, and inadequate approaches for evaluating models.

In this study we used routinely collected data from a single National Health Service (NHS) hospital trust to create and evaluate ML models to predict and forecast outcomes in hospital admissions from patients diagnosed with CAP (30-day mortality and CURB65 at the third day of hospitalisation). Here we present the results of our best performing models using different pre-processing approaches and both single time-point and time series data. We then compare our results with other comparable models and identify the strengths and weaknesses of our approach.

## Methods

### Data sources

Ethical approval for the study was obtained from the regional NHS Research Ethics Committee (IRAS ID: 266731). Anonymised data was extracted from electronic health record (EHR) admissions of patients diagnosed with CAP at the University Hospitals of Leicester (UHL) between 2016, and February 2020. Data collected during the COVID19 pandemic (after February 2020) was excluded. Data was extracted by the National Institute of Health Research (NIHR) Leicester Biomedical Research Centre IT team. CAP patients were identified using appropriate International Classification of Diseases, tenth Revision (ICD-10) codes and a previously described approach [20]. The ICD-10 codes are listed in supplementary document A. Patients without immunosuppressive conditions (e.g., AIDS, or positive for HIV) and those aged under 16 were excluded.

### Data pre-processing

Data extracted consisted of information from admissions, medical history, vital signs and blood tests (34 variables). It included eight static demographics variables (e.g., age at admission, ethnicity, and mortality) and 26 time series variables (e.g., respiratory rate, blood pressure). Where required, binary mental confusion was determined by deriving it from other scores (e.g., Early Warning Score). Charlson Comorbidity Index was calculated using previously reported weights [22]. Data was divided into two batches. Batch one (admissions from 2016-2018) was used to train models (training set). Batch two (admissions from 2019 – February 2020) was used to validate models during their development (test set). Data was pre-processed following the process in Figure 1 to produce linked tabular data sets (e.g., delimited comma-separated value files), merging tables, then filtering, cleaning, and interpolating. The same process was undertaken for both the training set and test set.

**Figure 1.**
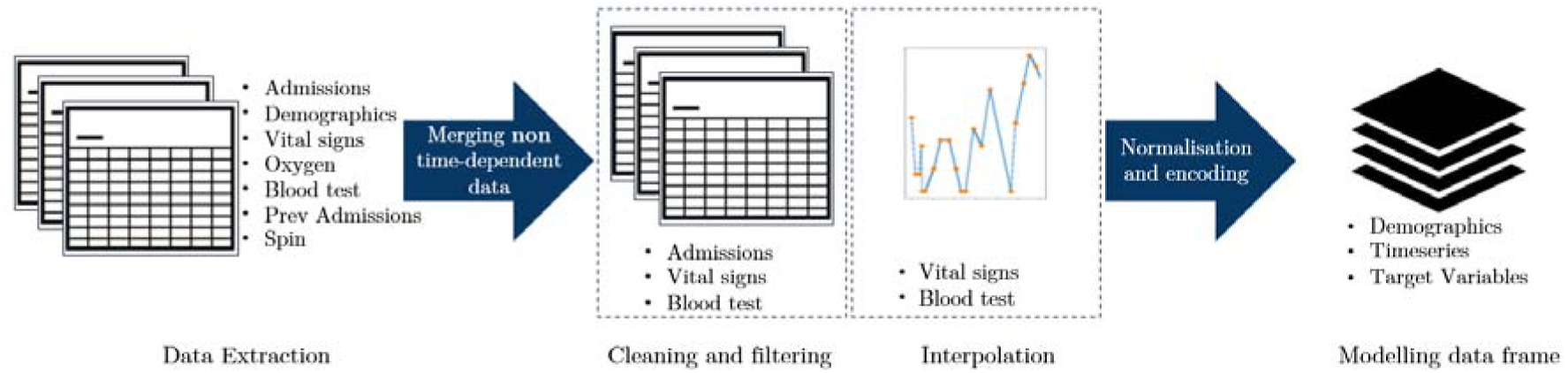
Process flow of information processing from UHL data extraction to the final dataset used for modelling.

Admissions were only included in the data sets if they met the following inclusion criteria: i) diagnosis with CAP (see above); ii) length of stay equal or longer than 3 days; iii) more than 6 records available for both vital signs and blood tests. Data cleaning consisted of bounding data within ranges of features. These ranges were defined based on clinical utility and are presented in supplementary document B). Only blood test variables which were collected in ≥70% of admissions were included.

Filtered-cleaned data from time series (vital signs and blood tests) was sparse and uneven, so interpolation was performed to derive values spaced at 30-minute intervals across 3 days. Interpolation took place between the first known value for every variable in each admission to the last. Where more than one value was present in a period, a mean value was calculated. Multiple interpolation methods (linear, polynomial, Akima spline, piecewise cubic Hermite interpolation polynomial and nearest neighbour) were evaluated in a random selected sample of 25% of the training data, and using cross-validation the method with the lowest mean square error (MSE) per variable was identified and subsequently used. The above methods were then used to interpolate the variable in both, training and test sets.

Time series data was then encoded using max-min scaling. The final time series dataset contained the interpolated time series of all included admissions comprising 144 readings per admission per vital sign, and blood test.

For data representing static time points, continuous variables were normalised. Target-encoder was used to encode categorical variables as described in [23], to reduce the impact of the difference between the order of magnitude in the variable, and to transform variables in values of a range between 0 and 1 preserving most of the predictive power of the original variables [23]. Every admission was considered independent, including re-admissions.

### Statistical analysis of independent variables

Differences across 34 variables between patients who were deceased and discharged were assessed. This analysis utilized the first value for each variable collected at admission, providing insights into the patient’s baseline condition at hospital arrival. Significance was tested for continuous variables via the t-test, for binary variables using the normal test, and for categorical variables using Kruskal-Wallis test, with a 95% significance level.

### Mortality modelling

Mortality was defined as a binary variable denoting death in-hospital or within 30-days of hospital discharge. Several modelling components were explored to determine their impact on model performance. These components included: collection time, stratification by age, data balance towards the target variable, and classifier model type. Distinct attributes of these components are presented in **Error! Not a valid bookmark self-reference.** with the rationale behind their selection in this paper. Time series are identified as one type of component, to support benchmarking against models built using non-time dependent data. All models used 34 variables, when models involved time-dependent data, they were combined with demographics / static variables (e.g., age at admission, ethnicity, etc) Non-time series hyper-parameters were fine-tuned using search grid method and 10-fold cross-validation for metrics calculation, hyper-parameters and values can be found at supplementary material E. Whilst time series classifiers (LSTM and GRU) were optimised using a Bayesian approach previously demonstrated to find optimal hyper-parameters [24–26], with 5-fold cross-validation. Models were evaluated using multiple metrics including the area under the receiver operating characteristics curve (AUROC), recall, accuracy, precision, and F1 score.

Hyper-parameter tuning used the search grid method and 10-fold cross-validation for SVM, Random Forest and XGBoost classifiers. Whilst time series classifiers (LSTM and GRU) were fine-tuned by applying a Bayesian optimisation approach to find optimal hyper-parameters in artificial neural networks [24–26], with 5-fold cross-validation.

### CAP severity forecasting

Severity was represented by URB65, a variation of the CURB65 score excluding confusion (not included in the initial extraction but calculated from EWS). This was derived by forecasting required vital signs individually (blood urea level, respiratory rate, and blood pressure) on the third day and then calculating a score as per CURB65 [27]. The predicted URB65 score (severity in a scale of 0-4 scoring 1 point for each when the following are present: urea >7 mmol/L; respiratory rate ≥ 30/minute, systolic blood pressure <90 mmHg and/or diastolic blood pressure ≤ 0 mmHg; and age ≥ 65 years [27]) was calculated at the end of the third day of hospitalisation using the predictions from the models described below.

Two components were built to forecast vital signs. First, a max-min scaling component with global and local attributes for time series transformation. The former used the max-min reference from each of the individual time series, whereas the latter used max-min reference values from across the entire data set. Second, ML models components (GRU and LSTM) predicted the sequence of the last day of hospitalisation (48 values for each variable), using a many-to-many methodology [15, 28] where neural networks are designed which sequence the first two days (96 values), and predict the sequence of each variable at the third day (48 values) of hospitalisation. Hyper-parameters were tuned using Bayesian optimisation [25]. Model performance was assessed using root mean squared error (RMSE). General MSE refers to the average MSE of the series predicted, while the last MSE represents the last value at the third day of hospitalisation. The final URB65 results is compared using the confusion matrix, the global accuracy, and the accuracy per URB65 class.

The predicted URB65 score (severity in a scale of 0-4 scoring 1 point for each when the following are present: urea >7 mmol/L; respiratory rate ≥ 30/minute, systolic blood pressure <90 mmHg and/or diastolic blood pressure ≤ 0 mmHg; and age ≥ 65 years [27]) was calculated at the end of the third day of hospitalisation using the predictions from the models of the involved vital signs. The final URB65 results is compared using the confusion matrix, the global accuracy, the accuracy per URB65 class and MatchR, a parallel for recall in multiclass classification, assessing the accuracy of prediction for each class.

### Tools and statistical frameworks

All data processing and analytical work was done using Python 3.8 [29]. Preprocessing, and cleaning used Pandas version 2.0.3 [30]. Interpolation, modelling and statistical analysis used statsmodels version 0.13.2 [31], scikit-learn modelling [32], Keras 2.0 machine learning framework [33], Tensorflow version 2.10.0 [34] and Bayesian-optimisation [35] packages. Scripts are available at a GitHub repository in supplementary document C. This study used the ALICE High Performance Computing facility at the University of Leicester.

## Results

### Descriptive statistics

A summary of the cleaning and filtering process (inclusion and exclusion of admissions) is presented in Figure 2 for each of the datasets. The training set (A) comprised 24,720 admissions (18,104 patients). After pre-processing, 9,390 admissions were included (8,243 patients). Similarly, the test set (B) contained 24,720 admissions (20,088 patients) which were narrowed down to 7,892 admissions (7,236 patients).

**Figure 2.**
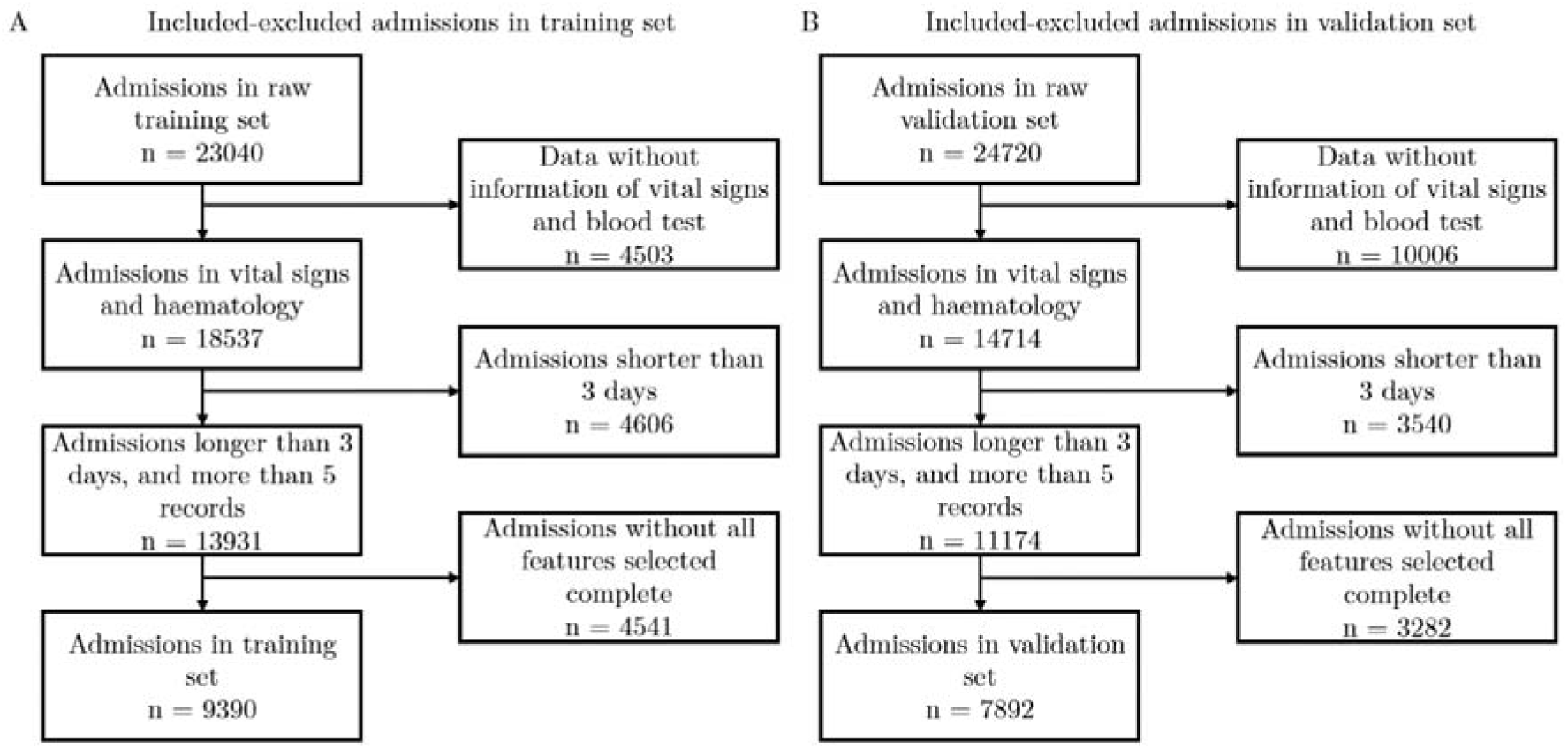
Flow chart showing inclusion and exclusion criteria of study

**Figure 3.**
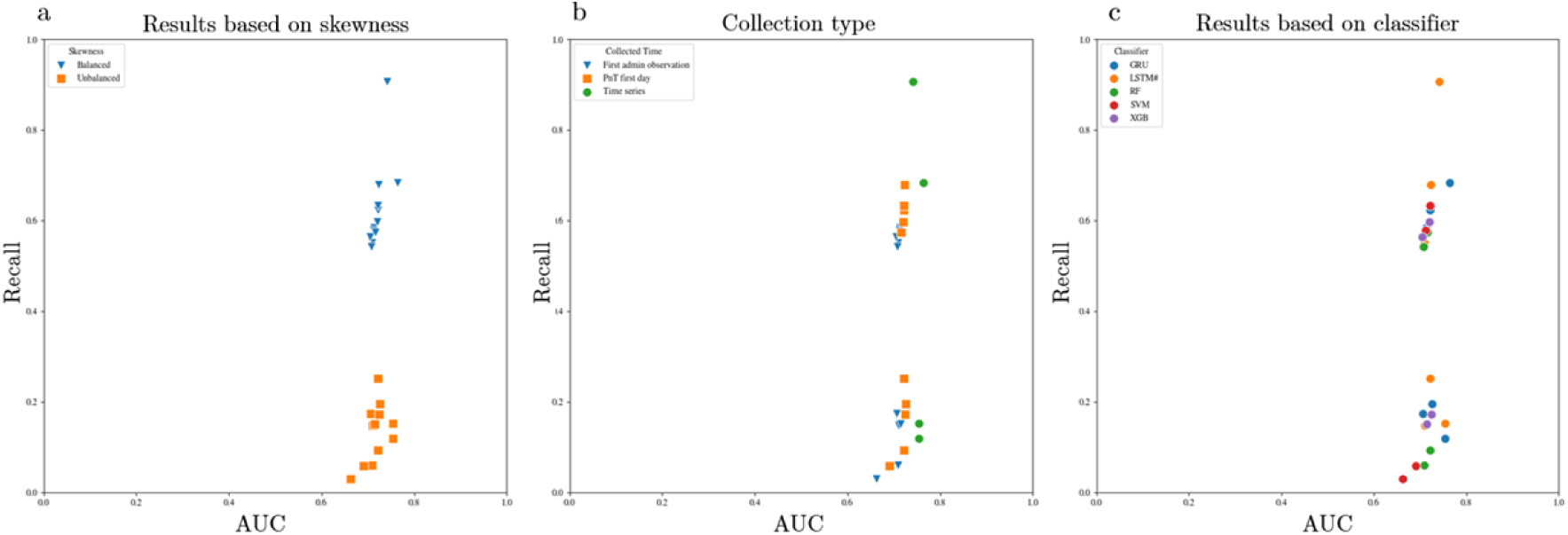
Results of CAP admissions mortality prediction models shown using AUC against recall. A) impact of a balancing dataset for training purposes in model results. B) All models developed and tested, divided by collected time component. C) Performance by classifier

Table 2 shows that the demographics of training and test sets were similar in nature. From the 9390 admissions that formed the training set 22% (2024) died in hospital or within 30 days of discharge, 48% (4553) were women, and 80% (7524) were recorded as white British with a median age of 78 (IQR 66 - 85), and a median Charlson comorbidity score of 1 (IQR 0 - 3). The test set included 7892 admissions in which 25% (1944) died in hospital or within 30 days of discharge, 45% (3541) were women, 72% (5667) were white British, with a median age of 74 (IQR 61 - 83) and a median Charlson comorbidity score of 2 (IQR 0 - 3).

**Table 1.** Components of modelling for comparing the effect of different attributes on CAP outcome prediction.

| Component | Type | Attribute | Rationale for selection |
| --- | --- | --- | --- |
| Data time range | Domain specific | • First observation at admission<br>• Both peak (highest) and trough (lowest) observations collected during the first day of admission (PnT)<br>• Time series across the first three days of hospitalisation | Clinical processes consider information in different instances of the admission that might bring better performance to the models. |
| Patients' stratification | Domain specific | • Age (over and under 65)<br>• No stratification | Identifying if tiered model approach may better predict outcome |
| Data balance of target variable | Data specific | • Unbalanced<br>• Balanced | Specific to this type of data where around 20% of the patients admitted did not survive. Balancing was achieved by random sampling of discharged admissions. |
| Classifier | Data specific | • Support Vector Machine (SVM)<br>• Random Forest (RF)<br>• XGBoost<br>• LSTM<br>• GRU | As an exploratory study the intention is to investigate what performance is achieved from different training models. A mix of models that considered time series and static variables were selected for this study. |

**Table 2.**
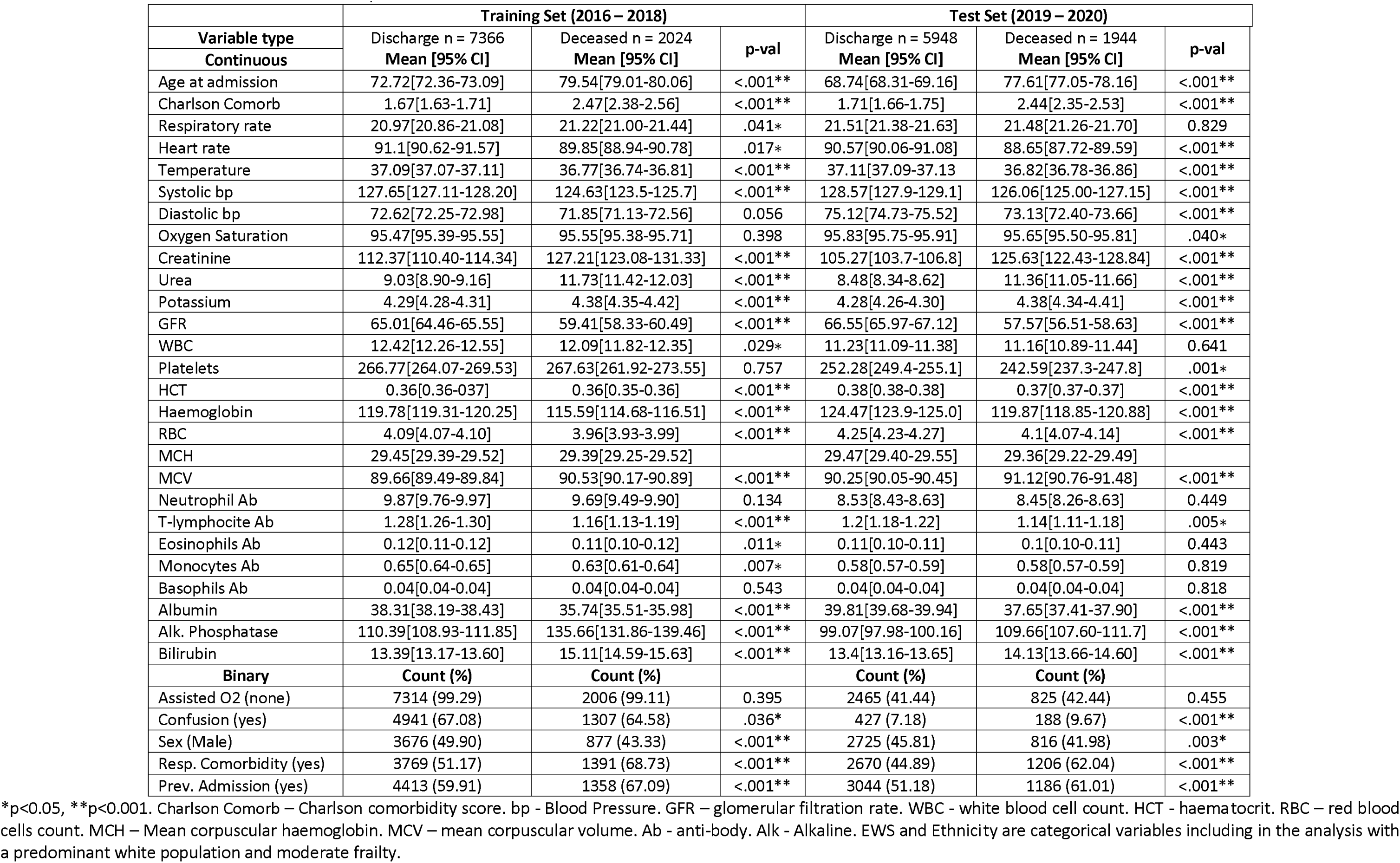
Baseline characteristics of admissions of patients with CAP for training and test data sets. Statistical comparison between admissions where patients were discharged and those who diseased.

|  | Training Set (2016 – 2018) |  |  | Test Set (2019 – 2020) |  |  |
| --- | --- | --- | --- | --- | --- | --- |
| Variable type | Discharge n = 7366 | Deceased n = 2024 | p-val | Discharge n = 5948 | Deceased n = 1944 | p-val |
| Continuous | Mean [95% CI] | Mean [95% CI] |  | Mean [95% CI] | Mean [95% CI] |  |
| Age at admission | 72.72[72.36-73.09] | 79.54[79.01-80.06] | <.001** | 68.74[68.31-69.16] | 77.61[77.05-78.16] | <.001** |
| Charlson Comorb | 1.67[1.63-1.71] | 2.47[2.38-2.56] | <.001** | 1.71[1.66-1.75] | 2.44[2.35-2.53] | <.001** |
| Respiratory rate | 20.97[20.86-21.08] | 21.22[21.00-21.44] | .041* | 21.51[21.38-21.63] | 21.48[21.26-21.70] | 0.829 |
| Heart rate | 91.1[90.62-91.57] | 89.85[88.94-90.78] | .017* | 90.57[90.06-91.08] | 88.65[87.72-89.59] | <.001** |
| Temperature | 37.09[37.07-37.11] | 36.77[36.74-36.81] | <.001** | 37.11[37.09-37.13] | 36.82[36.78-36.86] | <.001** |
| Systolic bp | 127.65[127.11-128.20] | 124.63[123.5-125.7] | <.001** | 128.57[127.9-129.1] | 126.06[125.00-127.15] | <.001** |
| Diastolic bp | 72.62[72.25-72.98] | 71.85[71.13-72.56] | 0.056 | 75.12[74.73-75.52] | 73.13[72.40-73.66] | <.001** |
| Oxygen Saturation | 95.47[95.39-95.55] | 95.55[95.38-95.71] | 0.398 | 95.83[95.75-95.91] | 95.65[95.50-95.81] | .040* |
| Creatinine | 112.37[110.40-114.34] | 127.21[123.08-131.33] | <.001** | 105.27[103.7-106.8] | 125.63[122.43-128.84] | <.001** |
| Urea | 9.03[8.90-9.16] | 11.73[11.42-12.03] | <.001** | 8.48[8.34-8.62] | 11.36[11.05-11.66] | <.001** |
| Potassium | 4.29[4.28-4.31] | 4.38[4.35-4.42] | <.001** | 4.28[4.26-4.30] | 4.38[4.34-4.41] | <.001** |
| GFR | 65.01[64.46-65.55] | 59.41[58.33-60.49] | <.001** | 66.55[65.97-67.12] | 57.57[56.51-58.63] | <.001** |
| WBC | 12.42[12.26-12.55] | 12.09[11.82-12.35] | .029* | 11.23[11.09-11.38] | 11.16[10.89-11.44] | 0.641 |
| Platelets | 266.77[264.07-269.53] | 267.63[261.92-273.55] | 0.757 | 252.28[249.4-255.1] | 242.59[237.3-247.8] | .001* |
| HCT | 0.36[0.36-0.37] | 0.36[0.35-0.36] | <.001** | 0.38[0.38-0.38] | 0.37[0.37-0.37] | <.001** |
| Haemoglobin | 119.78[119.31-120.25] | 115.59[114.68-116.51] | <.001** | 124.47[123.9-125.0] | 119.87[118.85-120.88] | <.001** |
| RBC | 4.09[4.07-4.10] | 3.96[3.93-3.99] | <.001** | 4.25[4.23-4.27] | 4.1[4.07-4.14] | <.001** |
| MCH | 29.45[29.39-29.52] | 29.39[29.25-29.52] |  | 29.47[29.40-29.55] | 29.36[29.22-29.49] |  |
| MCV | 89.66[89.49-89.84] | 90.53[90.17-90.89] | <.001** | 90.25[90.05-90.45] | 91.12[90.76-91.48] | <.001** |
| Neutrophil Ab | 9.87[9.76-9.97] | 9.69[9.49-9.90] | 0.134 | 8.53[8.43-8.63] | 8.45[8.26-8.63] | 0.449 |
| T-lymphocyte Ab | 1.28[1.26-1.30] | 1.16[1.13-1.19] | <.001** | 1.2[1.18-1.22] | 1.14[1.11-1.18] | .005* |
| Eosinophils Ab | 0.12[0.11-0.12] | 0.11[0.10-0.12] | .011* | 0.11[0.10-0.11] | 0.1[0.10-0.11] | 0.443 |
| Monocytes Ab | 0.65[0.64-0.65] | 0.63[0.61-0.64] | .007* | 0.58[0.57-0.59] | 0.58[0.57-0.59] | 0.819 |
| Basophils Ab | 0.04[0.04-0.04] | 0.04[0.04-0.04] | 0.543 | 0.04[0.04-0.04] | 0.04[0.04-0.04] | 0.818 |
| Albumin | 38.31[38.19-38.43] | 35.74[35.51-35.98] | <.001** | 39.81[39.68-39.94] | 37.65[37.41-37.90] | <.001** |
| Alk. Phosphatase | 110.39[108.93-111.85] | 135.66[131.86-139.46] | <.001** | 99.07[97.98-100.16] | 109.66[107.60-111.7] | <.001** |
| Bilirubin | 13.39[13.17-13.60] | 15.11[14.59-15.63] | <.001** | 13.4[13.16-13.65] | 14.13[13.66-14.60] | <.001** |
| Binary | Count (%) | Count (%) |  | Count (%) | Count (%) |  |
| Assisted O2 (none) | 7314 (99.29) | 2006 (99.11) | 0.395 | 2465 (41.44) | 825 (42.44) | 0.455 |
| Confusion (yes) | 4941 (67.08) | 1307 (64.58) | .036* | 427 (7.18) | 188 (9.67) | <.001** |
| Sex (Male) | 3676 (49.90) | 877 (43.33) | <.001** | 2725 (45.81) | 816 (41.98) | .003* |
| Resp. Comorbidity (yes) | 3769 (51.17) | 1391 (68.73) | <.001** | 2670 (44.89) | 1206 (62.04) | <.001** |
| Prev. Admission (yes) | 4413 (59.91) | 1358 (67.09) | <.001** | 3044 (51.18) | 1186 (61.01) | <.001** |

\*p<0.05, \*\*p<0.001. Charlson Comorb – Charlson comorbidity score. bp - Blood Pressure. GFR – glomerular filtration rate. WBC - white blood cell count. HCT - haematocrit. RBC – red blood cells count. MCH – Mean corpuscular haemoglobin. MCV – mean corpuscular volume. Ab - anti-body. Alk - Alkaline. EWS and Ethnicity are categorical variables including in the analysis with a predominant white population and moderate frailty.

The comparison between discharged and deceased patients were identified, and showed significant differences (p<0.05) in key vital signs (temperature, blood pressure, heart rate) and common blood tests (creatinine, urea, cells blood counts, potassium, albumin, alkaline phosphate), with only mean corpuscular haemoglobin (MCH) and assisted O_2_ not showing significant differences.

### Predicting Mortality in CAP admissions

A total of 24 models were created using all training data (unstratified). Twenty for non-time dependent data corresponding to two types of data time collection (first value collected within the admission and peak and trough (PnT) values across three days), data balanced/unbalanced and five different classifiers. Four models were created using time series data, based on two different recurrent neural network-based classifiers and balanced/unbalanced data.

The performance metrics of the models are presented in Table 3. The best performing model used a recurrent neural network (RNN) based approach: a Gated Recurrent Unit (GRU) classifier trained using time series data, and a balanced data set (AUROC 0.77 and recall 0.69). This outperformed a Support Vector Machine (SVM) classifier (AUROC 0.72 and recall 0.63) and another RNN-based method, Long Short-Term Memory (LSTM) (AUROC 0.72 and recall 0.67) both of which used balanced data sets and PnT data for training. All models trained using balanced data produced higher recall than those trained using unbalanced data (maximum 0.25).

**Table 3.** Performance metrics of the 24 CAP admissions mortality prediction models ordered descending by best performance assessed by AUC and F1-score.

| Data collection time | Target variable balanced | Classifier | AUC | Recall | Precision | Accuracy | F1-score |
| --- | --- | --- | --- | --- | --- | --- | --- |
| Time series | Yes | GRU | 0.77 | 0.69 | 0.43 | 0.70 | 0.53 |
| Pnt | Yes | SVM | 0.72 | 0.63 | 0.40 | 0.68 | 0.49 |
| Pnt | Yes | LSTM | 0.72 | 0.67 | 0.38 | 0.65 | 0.49 |
| Pnt | Yes | GRU | 0.72 | 0.62 | 0.39 | 0.67 | 0.48 |
| Pnt | Yes | XGB | 0.72 | 0.59 | 0.40 | 0.68 | 0.48 |
| Pnt | Yes | RF | 0.71 | 0.57 | 0.40 | 0.68 | 0.47 |
| 1st | Yes | SVM | 0.71 | 0.57 | 0.40 | 0.68 | 0.47 |
| 1st | Yes | GRU | 0.71 | 0.58 | 0.39 | 0.68 | 0.47 |
| 1st | Yes | XGB | 0.70 | 0.56 | 0.39 | 0.68 | 0.46 |
| 1st | Yes | LSTM | 0.70 | 0.55 | 0.40 | 0.68 | 0.46 |
| Time series | Yes | LSTM | 0.74 | 0.91 | 0.31 | 0.48 | 0.46 |
| 1st | Yes | RF | 0.70 | 0.54 | 0.39 | 0.68 | 0.45 |
| Pnt | No | LSTM | 0.72 | 0.25 | 0.52 | 0.75 | 0.33 |
| Pnt | No | GRU | 0.72 | 0.19 | 0.56 | 0.76 | 0.29 |
| 1st | No | GRU | 0.70 | 0.17 | 0.54 | 0.76 | 0.26 |
| Pnt | No | XGB | 0.72 | 0.17 | 0.54 | 0.76 | 0.26 |
| 3days | No | LSTM | 0.75 | 0.15 | 0.66 | 0.77 | 0.24 |
| 1st | No | XGB | 0.71 | 0.15 | 0.57 | 0.76 | 0.23 |
| 1st | No | LSTM | 0.71 | 0.14 | 0.58 | 0.76 | 0.23 |
| 3days | No | GRU | 0.75 | 0.12 | 0.73 | 0.77 | 0.20 |
| Pnt | No | RF | 0.72 | 0.09 | 0.63 | 0.76 | 0.16 |
| 1st | No | RF | 0.70 | 0.06 | 0.66 | 0.76 | 0.11 |
| Pnt | No | SVM | 0.69 | 0.05 | 0.69 | 0.76 | 0.10 |
| 1st | No | SVM | 0.66 | 0.03 | 0.73 | 0.75 | 0.05 |

#### 1.1.1. Age Stratification

It is known that CAP patients admitted to hospital aged 65 and above have worse outcomes. To explore whether stratified models which incorporate this age boundary could improve performance, we created stratified models by training them on datasets divided into age groups (under 65 years old, and greater than or equal to 65 years old). For patients under 65, the training set contained 2196 admissions (252 deceased), and the test set 2533 admissions (306 deceased). This division did affect the balance of the classes in the data, with a mortality rate of 25% for patients 65 and above, which dropped to 9% for patients under 65 (compared to 21% for the overall dataset).

In total, 40 models were trained (20 per age group), using the same three approaches described in the previous section (time range, classifier type, and balanced). The results from the best performing models are presented in Table 4. For both patient groups, the top models utilised PnT data and balanced classes, but for patients ≥65 the most effective classifier was GRU (AUROC=0.68, recall=0.66, F1 score = 51) while for patients < 65, the most effective was an SVM classifier (AUROC = 0.71, recall = 0.46, F1 score = 0.33). However, the performance of the latter suffers from poor recall and F1 score. More complex RNN-based and SVM classifiers were shown to perform better than others.

**Table 4.** Age stratification metrics for the most important models Performance corresponds of a balance between the AUC and the F1-score which is compound by recall and precision.

| Age Group | Time range | Target variable balanced | Classifier | AUC | Recall | Precision | Accuracy | F1 score |
| --- | --- | --- | --- | --- | --- | --- | --- | --- |
| ≥ 65 | PnT | Yes | GRU | 0.68 | 0.66 | 0.42 | 0.61 | 0.51 |
|  | PnT | Yes | LSTM | 0.67 | 0.63 | 0.42 | 0.62 | 0.51 |
|  | PnT | Yes | SVM | 0.67 | 0.61 | 0.43 | 0.63 | 0.5 |
|  | PnT | Yes | ANN | 0.67 | 0.6 | 0.43 | 0.64 | 0.5 |
|  | PnT | Yes | XGB | 0.66 | 0.61 | 0.42 | 0.62 | 0.5 |
| < 65 | PnT | Yes | SVM | 0.71 | 0.46 | 0.26 | 0.76 | 0.33 |
|  | 1st | Yes | GRU | 0.70 | 0.48 | 0.25 | 0.77 | 0.33 |
|  | 1st | Yes | SVM | 0.71 | 0.44 | 0.25 | 0.78 | 0.33 |
|  | 1st | Yes | LR | 0.71 | 0.45 | 0.26 | 0.77 | 0.32 |
|  | 1st | Yes | XGB | 0.70 | 0.57 | 0.23 | 0.71 | 0.32 |

### 1.2. Forecasting CAP Severity

#### 1.2.1. URB65 variables forecasting

A forecasting approach was devised which utilised models that predicted each of the four vital signs over a three-day period (respiratory rate, systolic blood pressure, diastolic blood pressure, blood urea), where in two days (96 readings) were used to train a model to predict the third day (48 readings). The results obtained at the end of the third day i.e. the last value of the predicted sequence was used to predict a severity score for the third day (URB65).

Sixteen models were created using all processed admissions as training data (9390). Four models were created for each of the four vital signs, based on RNNs (LSTM and GRU) and two variations of the max-min scaling method (global or local). RNNs are designed to handle time series data better than other models and had also previously produced the best performing models. Performance was evaluated against all admissions in the test set (7892). Table 5 shows the root mean squared error (RMSE) for each vital sign, and shows that overall global scaling outperformed local max-min scaling. There was little difference in the results obtained from the two types of models (less than 0.05). The smallest RMSE was reported by the ‘blood urea’ model using global scaling (General RMSE = 0.08). RMSE based, global max-min transformation performed better than local max-min where general RMSEs show large increments for each of the symptoms involved in the URB65 calculation. Both types of blood pressure generated larger RMSE, in most of the cases >1, suggesting errors bigger than the max value used for scaling and leading to poor performance in these variables.

**Table 5.** results when forecasting variables involve URB65 General RMSE refers to the mean error of all the values predicted. Last RMSE refers to the RMSE of the last instance predicted. IQR is the interquartile range.

| Vital Sign | Scaling | Model | General RMSE [iqr] | Last RMSE [ iqr] |
| --- | --- | --- | --- | --- |
| Respiratory Rate | Global | GRU | 0.13 [0.03 - 0.76] | 0.03 [0.01 - 0.09] |
|  |  | LSTM | 0.12 [0.03 - 0.81] | 0.02 [0.01 - 0.08] |
|  | Local | GRU | 0.99 [0.22 - 3.26] | 0.16 [0.03 - 0.50] |
|  |  | LSTM | 0.98 [0.22 - 3.22] | 0.12 [0.02 - 0.43] |
| Systolic blood pressure | Global | GRU | 1.19 [0.26 - 3.74] | 0.43 [0.09 - 1.40] |
|  |  | LSTM | 1.14 [0.24 - 3.57] | 0.42 [0.08 - 1.41] |
|  | Local | GRU | 4.47 [0.99 - 12.7] | 1.66 [0.38 - 4.79] |
|  |  | LSTM | 4.40 [1.01 - 12.6] | 1.70 [0.39 - 4.80] |
| Diastolic blood pressure | Global | GRU | 1.07 [0.23 - 3.35] | 0.39 [0.08 - 1.24] |
|  |  | LSTM | 0.91 [0.19 - 2.89] | 0.34 [0.07 - 1.14] |
|  | Local | GRU | 2.64 [0.59 - 7.89] | 0.84 [0.18 - 2.55] |
|  |  | LSTM | 2.58 [0.57 - 7.74] | 0.86 [0.19 - 2.59] |
| Blood urea | Global | GRU | 0.08 [0.01 - 0.28] | 0.02 [0.00 - 0.07] |
|  |  | LSTM | 0.08 [0.01 - 0.31] | 0.02 [0.00 - 0.07] |
|  | Local | GRU | 1.17 [0.39 - 2.49] | 0.56 [0.18 - 1.29] |
|  |  | LSTM | 1.19 [0.41 - 2.52] | 0.57 [0.18 - 1.29] |

**Table 6.** Overall results of calculating severity with vital signs forecasts.

| Scaling | Overall Accuracy | Metrics | URB65 (n = 7236) |  |  |  |  |
| --- | --- | --- | --- | --- | --- | --- | --- |
|  |  |  | 0<br>n=1320<br>(17%) | 1<br>n=2493<br>(31%) | 2<br>n=3350<br>(42%) | 3<br>n=721<br>(9%) | 4<br>n=8<br>(0.01%) |
| Local GRU | 0.85 | Accuracy | 0.95 | 0.9 | 0.87 | 0.36 | 0.12 |
|  |  | Match R | 0.89 | 0.88 | 0.84 | 0.62 | 1 |
| Local LSTM | 0.85 | Accuracy | 0.97 | 0.9 | 0.88 | 0.29 | 0.12 |
|  |  | Match R | 0.89 | 0.87 | 0.83 | 0.65 | 1 |
| Global GRU | 0.78 | Accuracy | 0.98 | 0.83 | 0.78 | 0.22 | 0 |
|  |  | Match R | 0.78 | 0.75 | 0.82 | 0.62 | 0 |
| Global LSTM | 0.79 | Accuracy | 0.98 | 0.84 | 0.8 | 0.19 | 0 |
|  |  | Match R | 0.79 | 0.76 | 0.81 | 0.61 | 0 |

Results from the last RMSE were generally lower than the average general RMSE suggesting that the main differences might occur within the time series rather than at its last point, indicating that the global approach for respiratory rate and blood urea is a good approach to identify the state at the end of the day for each admission.

Figure 4 and Figure 5 show the patterns predicted using the GRU and LSTM models, respectively. Funnel shapes can be seen for all vital signs but blood urea when using local scaling, while global scaling produced more fluctuations in the trajectory of the prediction. There are also differences between LSTM and GRU models, with the former showing less fluctuation than the latter, and stabilising more rapidly. Additionally, values that are close to minimum values of the scale (e.g., values below 0.2) were predicted flat, with little increments regardless of the model.

**Figure 4.**
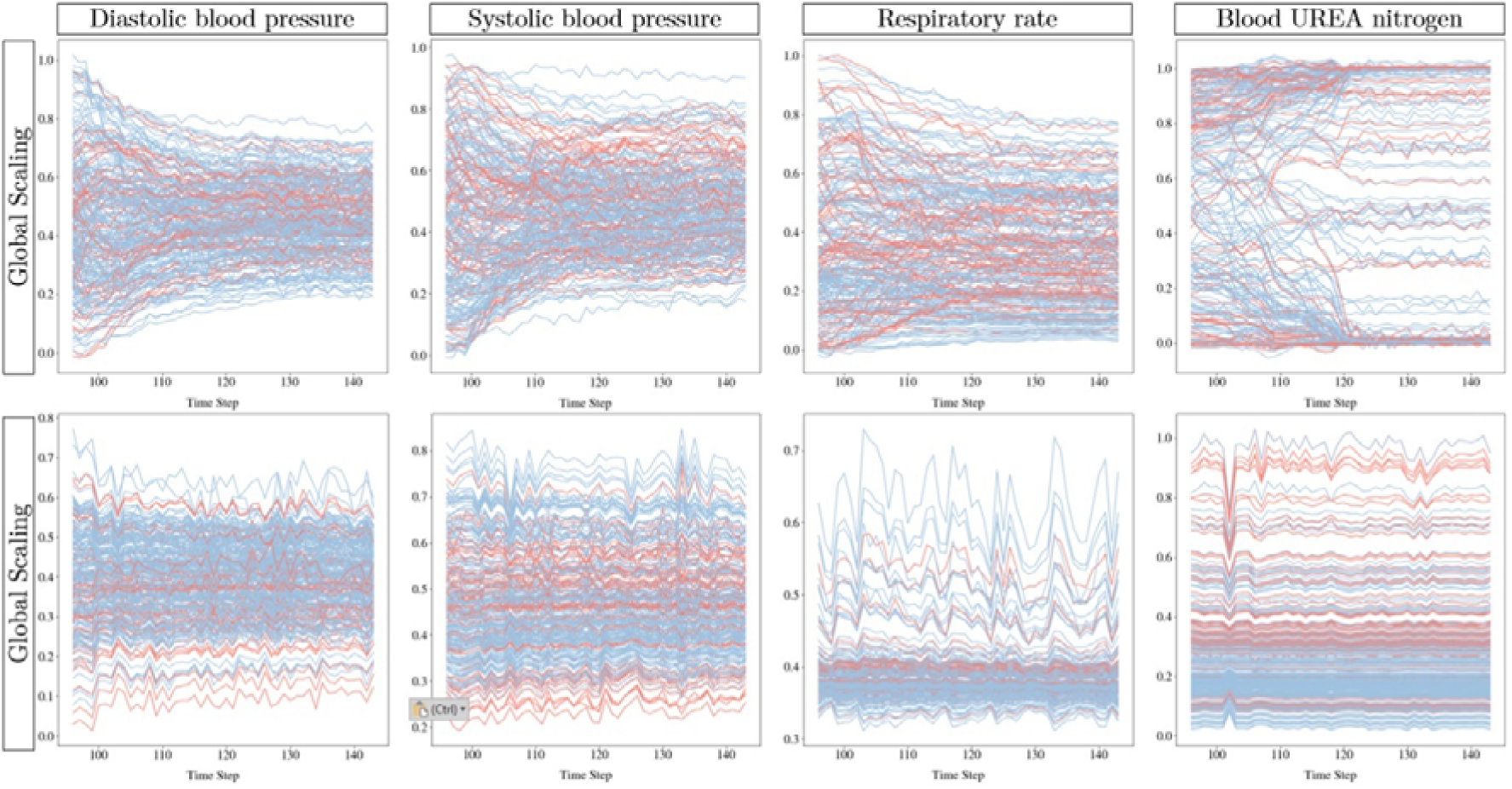
Predicted segment of the series by scaling characteristic and vital sign using GRU model. Plots of the last day of admissions (time points 97 to 144). Local scaling models tend to converge to a same point, while global models present bigger fluctuation, showing a more specific pattern in both. Blue lines represent admissions of patients that were discharge, and red of deceased admissions, showing no particular difference in the pattern.

**Figure 5.**
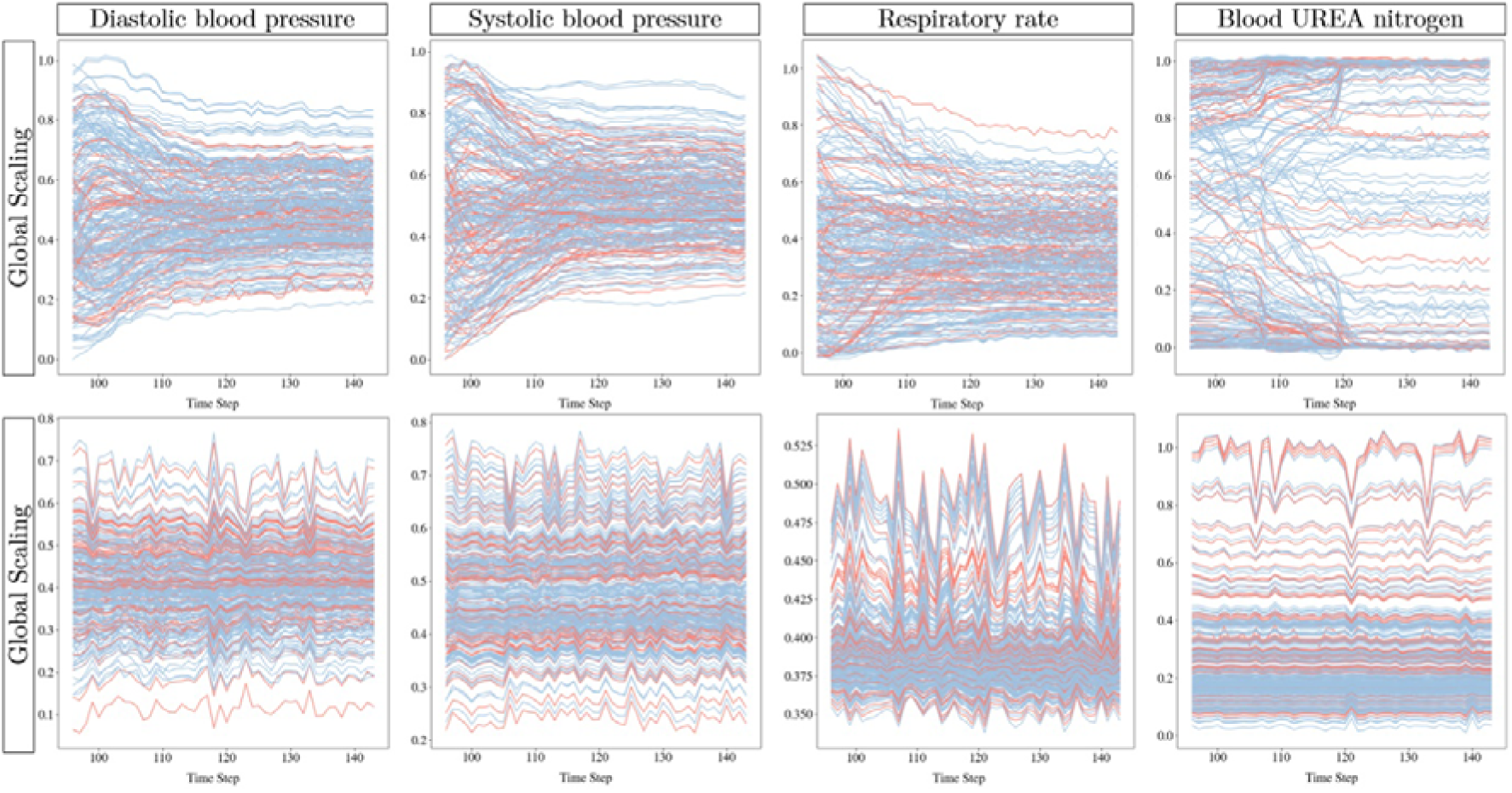
Predicted segment of the series by scaling characteristic and vital sign using LSTM model. Plots of the last day of admissions (time points 97 to 144). Local scaling models tend to converge to a same point, while global models present bigger fluctuation, showing a more specific pattern in both. Blue lines represent admissions of patients that were discharge, and red of deceased admissions, showing no particular difference in the pattern.

### Predicting CAP severity

The URB65 at the end of the third day was calculated for each admission using the real vital sign values, and compared with predicted values.

Predicted values of URB65 were obtained using the vital signs models presented above. All models were better at predicting low and medium severity classes (URB65 = 0,1,2), than high severity classes (URB65 = 3,4) and the accuracy gradually reduces across the classes, possibly a result of less instances in these latter classes. Interestingly, the CAP severity prediction accuracy was higher in models that used the local scaling method - with the highest total accuracy achieved by both GRU and LSTM (0.85). However, these models show differences in the distribution of predictions across their confusion matrices (Figure 6).

**Figure 6.**
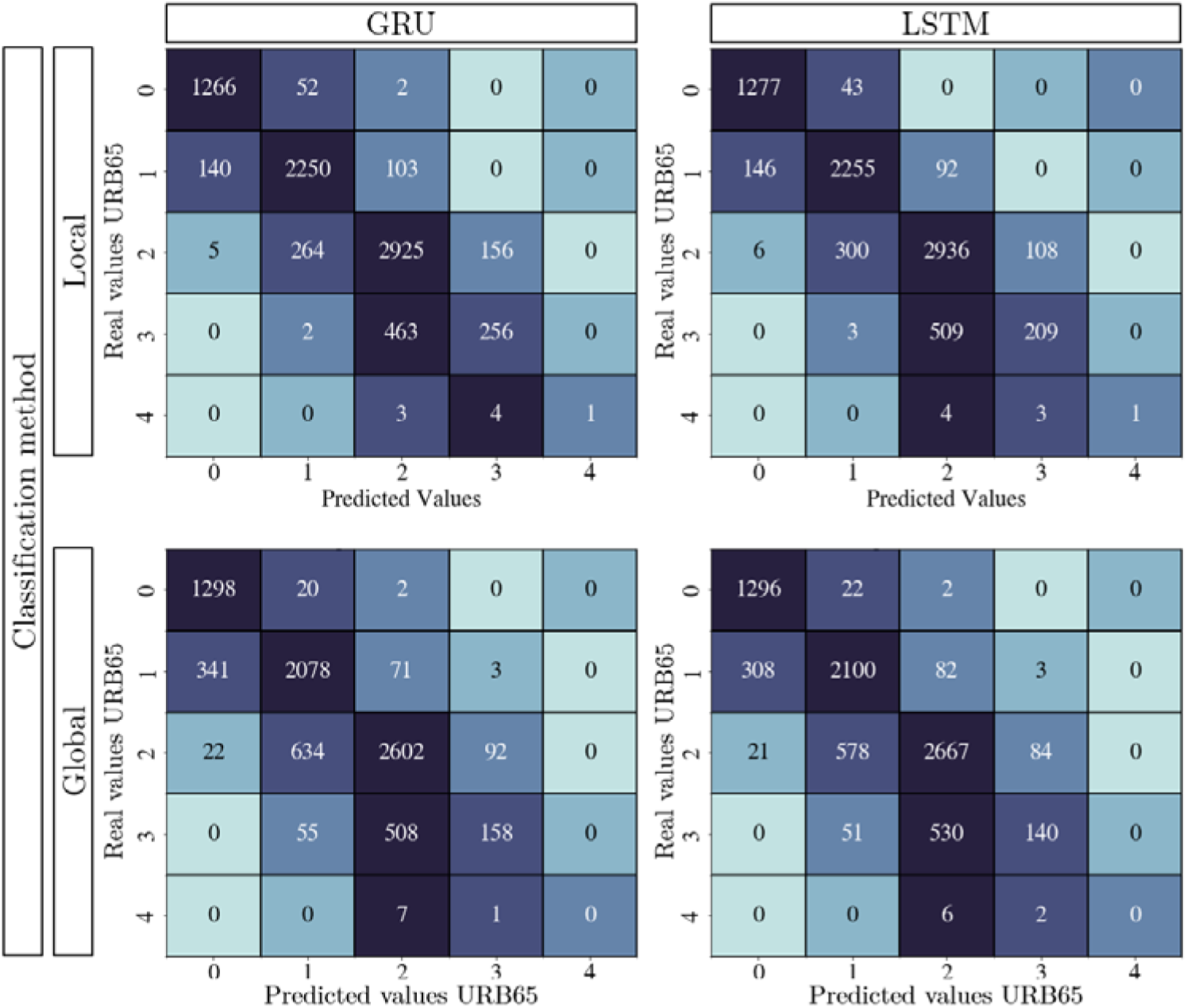
Comparison of URB65 by scaling and models

URB65 prediction of vital signs predicted using global scaling presented the same pattern in the distribution of predictions. However, it was limited due to a lack of admissions data, it was not possible to create a model to support predictions of URB65 = 4.

## Discussion

This study appears to be the first attempt to use the complete time series of vital signs and blood tests together to predict CAP outcome using ML models. We showed the potential of several ML-based approaches to predict and forecast CAP outcome (mortality and severity as URB65) using non-time dependent and time dependent routinely collected health data. Recurrent neural networks presented as the most promising approaches for predicting patient mortality from time series data, but there also appears to be potential for simpler classifiers based on non-time dependent data. Furthermore, we explored different approaches for developing models and pre-processing data to optimise our results.

Our main findings show that using timeseries (with imputed values) and balancing the data produced models with better performance than those produced using data representing single time-points. Consequently, the best model corresponded to a GRU classifier trained on such data. This latter observation could be explained through the sensitivity of ML model for dominant patterns of dominant classes [28]. This result may be significant for other studies where classes are regularly highly unbalanced, which is common in medical domain and often unaddressed [36–39].

Interestingly, from the classifiers used, SVM showed a slightly better performance when processing single time-points than other classifiers. Surprisingly, results from age stratified models showed no improvement when compared to non-stratified models, despite evidence of greater CAP severity in the older population [6].

Compared to other studies (Table 7), our model benefits from using data that is routinely collected, which facilitates its validation, and potential implementation. Our model was also designed to align with current guidelines for CAP management of moderate and high severity patients [9–11] who require hospital admission. This means that it can make use of data from hospitals across three days of admission, evaluating disease’s progression.

**Table 7.**
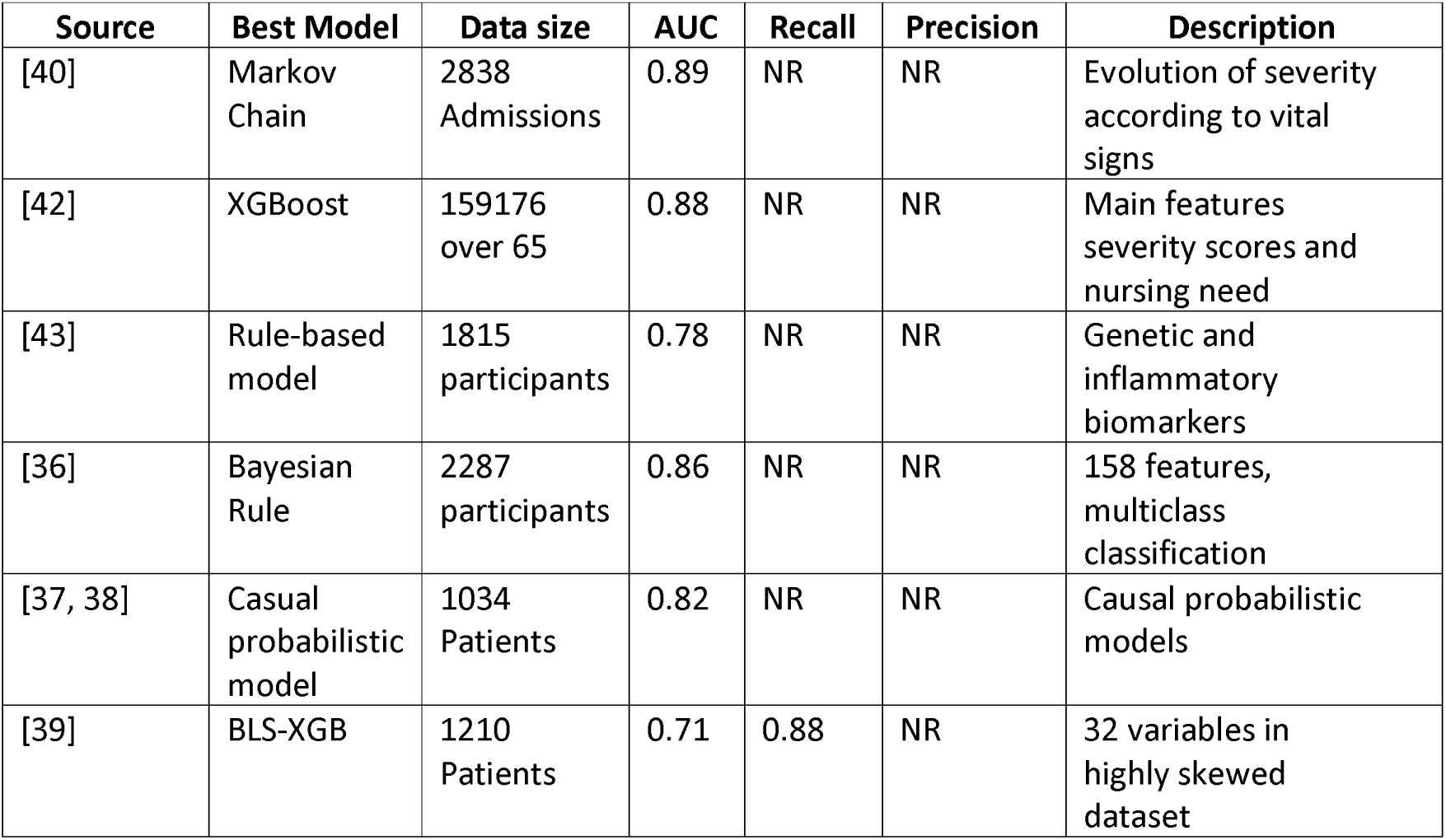
Studies of implementing ML models for CAP admission mortality prediction.

| Source | Best Model | Data size | AUC | Recall | Precision | Description |
| --- | --- | --- | --- | --- | --- | --- |
| [40] | Markov Chain | 2838 Admissions | 0.89 | NR | NR | Evolution of severity according to vital signs |
| [42] | XGBoost | 159176 over 65 | 0.88 | NR | NR | Main features severity scores and nursing need |
| [43] | Rule-based model | 1815 participants | 0.78 | NR | NR | Genetic and inflammatory biomarkers |
| [36] | Bayesian Rule | 2287 participants | 0.86 | NR | NR | 158 features, multiclass classification |
| [37, 38] | Casual probabilistic model | 1034 Patients | 0.82 | NR | NR | Causal probabilistic models |
| [39] | BLS-XGB | 1210 Patients | 0.71 | 0.88 | NR | 32 variables in highly skewed dataset |

Others similar modelling studies have not used entire time series and have relied on data collected at admissions [37–39, 42]. The exceptions to this rule [40] evaluated changes in patient states over time rather than analysing specific values within admissions. However, they have not proposed the use of time series, since a big challenge in analysing time series data from routinely collected data is that they often are incomplete and sparse. Most of these studies also only reported one metric (AUC), making it difficult to identify potential biases and issues with performance. Moreover, [36–38] reported results over data that was unbalanced (11%, 4%, and 10% mortality, respectively) with no discussion of the impact of it on their results. [40] based their model on the progression of the disease: using the SOFA score to represent CAP severity, although this metric is not specific to CAP. Finally, [39] trained and compared classifier models using variables that would not be available at admission (e.g., treatment received) which seems unfeasible. All this is evidence of gaps in the field that our study aimed to addressed.

We utilised a novel two-stage method for predicting URB65 – a variation of CURB65 score (excluding mental confusion) – using ML-based vital sign forecasting models.

Predictions from models built using locally scaled data produced more accurate CAP severity predictions than their globally scaled equivalents (Figure 4 and Figure 5), in spite of the smaller error in global forecasts we surmise that prediction of overall trajectory is more important than prediction of specific variability across patient’s time series. This is also evidenced by models based on local-scaling models identifying more critical patients, and producing a score closer to the actual value than those based on global-scaling.

Our most promising results were obtained using GRU and LSTM, indicating that these are appropriate models when considering time series for CAP management. Nevertheless, there may be additional approaches such as Gaussian process or generative adversarial models (GAN) which warrant investigation as these have shown promising results in adapting to different unstructured data or generate data under specific contexts [44]. The accuracy of the models suggested that this two-tiered approach could be a good option for forecasting CAP severity.

The work presented here is preliminary research to explore the potential of time series data in ML-based models and it is important to acknowledge certain limitations and caveats. Data used to train and test models was collected from a single hospital site over a specific period of time, which while representing a diverse population still contains a limited number of individuals, and any further development of these ML models would require refinement and validation across larger populations to provide an appropriate evidence base. we used the most recent data in our dataset for testing, to confirm models worked when potential changes in management process were considered [15]. As they stand, models were also designed according to the threshold for non-critical patients to have been discharged [45], and therefore would only be useful for patients who spend more than 3 days in hospital, although different time horizons can be explored. Additionally, several factors which could influence a patient’s progression could not be included due to lack of data collection or difficulties in obtaining the data. This included detailed clinical history, the size of lung infiltrates (accumulation of liquid in the lungs visible through X-rays), the patient’s smoking status, dietary factors, etc.

## 2. Conclusions

Our results show that use of AI models built using routinely collected time-series data could improve CAP outcome prediction in some cases. The use of balanced data sets also showed the importance of this approach when creating reliable models. The use of time series data for predictive modelling in medical settings is still under explored and has clear potential for improving management of CAP and other medical conditions.

## Supporting information

ICD-10 codes included in the research

Ranges of clinical utility used for the study

GitHub link for scripts

Results by characteristics age stratification

hyper-parameters and values of models implemented

## List of Abbreviations

AUROC/AUC: Area under the receiver’s operating characteristic curve
BRC: Biomedical research centre
CAP: Community-acquired pneumonia
EHR: Electronic health records
eObs: Electronic observations
EOS: Eosinophils
EWS: Early warning score
GFR: Glomerular filtration rate
GRU: Gated recurrent unit
HCT: Haematocrit
ICD-10: International classification of diseases version 10
IQR: Interquartile range
LSTM: Long-Short term memory
MCH: Mean corpuscular haemoglobin
MCV: Mean corpuscular volume
ML: Machine learning
RMSE: Root-mean-squared error
NHS: National health service
PnT: Peak and Trough
RBC: Red blood cells
RF: Random forest
RNN: Recurrent neural network
ROC: Receiver’s operating characteristic
SVM: Support vector machines
UHL: University hospitals of leicester
WBC: White blood cells
XGB: Extreme gradient boosting

## Declarations

## Clinical trial number

Not applicable

## Ethics approval and consent to participate

This study was approved by the NHS Research Ethics Committee (ref. 20/WM/0144) under the IRAS ID 266731. Prior to analysis, all patient data were anonymized to protect privacy.

## Consent for publication

Not Applicable

### Availability of data and materials Competing interests

*The authors declare no competing interests*.

## Funding

This project was co-funded by the NIHR Leicester Biomedical Research Centre, the University of Leicester and Minciencias Colombia (Colombian Ministry of Science, Technology & Innovation).

### Authors’ contributions

Daniel Lozano-Rojas designed the study, developed the models, carried the analysis out, and wrote the first draft of the manuscript. Robert C. Free and Gerrit Woltmann conceived, supervised the study. Robert C. Free extracted the data, and contributed to the design, analysis and drafting the manuscript. Matthew Richardson and Gerrit Woltmann provided intellectual contributions and revised the manuscript. All authors contributed to manuscript revision, read, and approved the submission.

## Data Availability

The data that support the findings of this study are available from the authors but restrictions apply to the availability of these data, which were used under REC reference number 20/WM/0144, and so are not publicly available. Data are, however, available from the authors upon reasonable request and pertinent permissions and amendments of the responsible research committee.

## Acknowledgements

The research was carried out at the National Institute for Health and Care Research (NIHR) Leicester Biomedical Research Centre (BRC). This research used the ALICE High Performance Computing facility at the University of Leicester.

## References

[1] Institute for Health Metrics and Evaluation (IHME). Global Burden of Disease 2021: Findings from the GBD 2021 Study. Seattle,WA:IHME; 2024.

[2] GBD 2021 Lower Respiratory Infections and Antimicrobial Resistance Collaborators. Global, regional, and national incidence and mortality burden of non-COVID lower respiratory infections and aetiologies, 1990–2021: a systematic analysis from the Global Burden of Disease Study 2021. The Lancet Infectious Diseases. 2024.

[3] Cilloniz C, Torres A. What’s Next in Pneumonia? Archivos de Bronconeumologia. 2022;58:1–15.

[4] Torres A, Cilloniz C, Niederman MS, Menendez R, Chalmers J, Wunderink RG, et al. Pneumonia. Nature Reviews Disease Primers. 2021;7(1):25.

[5] Chalmers J, Campling J, Ellsbury G, Hawkey PM, Madhava H, Slack M. Community-Acquired Pneumonia in the United Kingdom: A Call to Action. Pneumonia. 2017;9(1):1–6.

[6] Cilloniz C, Dominedo C, Pericas JM, Rodriguez-Hurtado D, Torres A. Community-Acquired Pneumonia in Critically Ill Very Old Patients: A Growing Problem. European Respiratory Review. 2020;29(155):1–15.

[7] NHS Digital (2024). Hospital Admitted Patient Care Activity, 2023-24: Diagnosis. Hospital Episode Statistics; 2024.

[8] Campling J, Wright HF, Hall GC, Mugwagwa T, Vyse A, Mendes D, et al. Hospitalization costs of adult community-acquired pneumonia in England. Journal of medical economics. 2022;25:912–8.

[9] National Institute for Health and Care Excellence (NICE). Pneumonia (community-acquired): antimicrobial prescribing. NICE guideline [NG138]; 2019 Reviwed 2022.

[10] Woltmann G. Pneumonia/LRTI guidance for antibiotic prescribing (2021). NHS University Hospitals of Leicester; 2022

[11] Musa D. Clinical Guideline for the Management of Community Acquired Pneumonia (CAP): Emergency Department and Acute Medicine. NHS Norfolk and Norwich University Hospital(2024); 2024.

[12] Zaki HA, Hamdi Alkahlout B, Shaban E, Mohamed EH, Basharat K, Elsayed WAE, et al. The Battle of the Pneumonia Predictors: A Comprehensive Meta-Analysis Comparing the Pneumonia Severity Index (PSI) and the CURB-65 Score in Predicting Mortality and the Need for ICU Support. Cureus. 2023;7:29–15.

[13] Lozano-Rojas D, Free RC, McEwan AA, Woltmann G. A Systematic Literature Review of Machine Learning Applications for Community-Acquired Pneumonia. In: Proceedings of 2021 International Conference on Medical Imaging and Computer-Aided Diagnosis (MICAD 2021). vol. 784. Springer Singapore; 2022. p. 292-301.

[14] Hurbans R. Grokking Artificial Intelligence Algorithms. New York: Manning Publications; 2020.

[15] Chollet F. Deep Learning with Python. Manning Publications; 2018.

[16] Varshni D, Thakral K, Agarwal L, Nijhawan R, Mittal A. Pneumonia Detection Using CNN Based Feature Extraction. Proceedings of 2019 3^rd^ IEEE International Conference on Electrical, Computer and Communication Technologies, ICECCT 2019. 2019:1-7.

[17] Wang T, Cai M, Ouyang X, Cao Z, Cai T, Tan X, et al. Anomaly Detection Based on Convex Analysis: A Survey. Frontiers in Physics. 2022 Apr;10:873848.

[18] Chumbita M, Cilloniz C, Puerta-Alcalde P, Moreno-Garcıa E, Sanjuan G, Garcia-Pouton N, et al. Can Artificial Intelligence Improve the Management of Pneumonia. Journal of Clinical Medicine. 2020;9(1):248.

[19] Gilvary C, Madhukar N, Elkhader J, Elemento O. The Missing Pieces of Artificial Intelligence in Medicine. Trends in Pharmacological Sciences. 2019;40(8):555–64.

[20] Free RC, Richardson M, Pillay C, Hawkes K, Skeemer J, Broughton R, et al. Specialist Pneumonia Intervention Nurse Service Improves Pneumonia Care and Outcome. BMJ Open Respiratory Research. 2021 Aug;8(1):e000863.

[21] Royal College of Physicians. National Early Warning Score National Early Warning Score (NEWS) 2: Standardising the Assessment of Acute-Illness Severity in the NHS. Updated report of a working party; London: RCP, 2017.

[22] Charlson ME, Pompei P, Ales KL, MacKenzie CR. A New Method of Classifying Prognostic Comorbidity in Longitudinal Studies: Development and Validation. Journal of Chronic Diseases. 1987;40(5):373–83.

[23] Micci-Barreca D. A Preprocessing Scheme for High-Cardinality Categorical Attributes in Classification and Prediction Problems. ACM SIGKDD Explorations Newsletter. 2001 Jul;3(1):27–32.

[24] Cabrera D, Guaman A, Zhang S, Cerrada M, Sanchez RV, Cevallos J, et al. Bayesian approach and time series dimensionality reduction to LSTM-based model-building for fault diagnosis of a reciprocating compressor. Neurocomputing. 2020;380:51–66.

[25] Nguyen HP, Liu J, Zio E. A long-term prediction approach based on long short-term memory neural networks with automatic parameter optimization by Tree-structured Parzen Estimator and applied to time series data of NPP steam generators. Applied Soft Computing. 2020;89:106116.

[26] Snoek J, Larochelle H, Adams RP. Practical Bayesian Optimization of Machine Learning Algorithms. In: Pereira F, Burges CJ, Bottou L, Weinberger KQ, editors. Advances in Neural Information Processing Systems. vol. 25. Curran Associates, Inc.; 2012.

[27] Capelastegui A, Espana PP, Quintana JM, Areitio I, Gorordo I, Egurrola M, et al. Validation of a Predictive Rule for the Management of Community-Acquired Pneumonia. European Respiratory Journal. 2006;27(1):151–7.

[28] Howard J, Gugger S, Chintala S. Deep Learning for Coders with Fastai and PyTorch: AI Applications without a PhD. First edition ed. O’Reilly Media, Inc; 2020.

[29] Van Rossum G, Drake FL. Python 3 Reference Manual. CreateSpace; 2009.

[30] pandas development team T. pandas-dev/pandas: Pandas. Zenodo; 2020. Available from: 10.5281/zenodo.3509134.

[31] Seabold S, Perktold J. statsmodels: Econometric and statistical modelling with python. In: 9th Python in Science Conference; 2010.

[32] Pedregosa F, Varoquaux G, Gramfort A, Michel V, Thirion B, Grisel O, et al. Scikit-learn: Machine Learning in Python. Journal of Machine Learning Research. 2011;12:2825–30.

[33] Chollet F, et al. Keras. GitHub; 2015. Available from: https://github.com/fchollet/keras.

[34] Abadi M, Agarwal A, Barham P, Brevdo E, Chen Z, Citro C, et al. TensorFlow: Large-Scale Machine Learning on Heterogeneous Systems; 2015. Software available from tensorflow.org. Available from: https://www.tensorflow.org/.

[35] Nogueira F. Bayesian Optimization: Open source constrained global optimization tool for Python; 2014. Available from: https://github.com/fmfn/BayesianOptimization.

[36] Visweswaran S, Cooper GF. Patient-Specific Models for Predicting the Outcomes of Patients with Community Acquired Pneumonia. AMIA Symposium. 2005;(March 1994):759–63.

[37] Cilloniz C, Ward L, Mogensen ML, Pericas JM, Mendez R, Gabarrus A, et al. Machine-Learning Model for Mortality Prediction in Patients with Community-acquired Pneumonia: Development and Validation Study. Chest. 2022 Jul.

[38] Ward L, Mogensen ML, Campos CC, Ceccato A, Gabarrus A, Amaro R, et al. A Machine-Learning Model for Prediction of Mortality among Patients with Community-Acquired Pneumonia. European Congress for Clinical Micrbiology and Infectious Diseases (ECCMID). 2019;779.

[39] Yuan J, Liu X, Wang WF, Zhang JJ. A Broad Learning System to Predict the 28-Day Mortality of Patients Hospitalized with Community-Acquired Pneumonia: A Case-Control Study. Computational and Mathematical Methods in Medicine. 2022;2022.

[40] Przybilla J, Ahnert P, Bogatsch H, Bloos F, Brunkhorst FM, Bauer M, et al. Markov State Modelling of Disease Courses and Mortality Risks of Patients with Community-Acquired Pneumonia. Journal of Clinical Medicine. 2020;9(2):393.

[41] Jones BE, Jones J, Bewick T, Lim WS, Aronsky D, Brown SM, et al. CURB-65 Pneumonia Severity Assessment Adapted for Electronic Decision Support. Chest. 2011 Jul;140(1):156–63.

[42] Shimizu S, Hara S, Fushimi K. PRS55 Predicting the risk of in-hospital mortality in adult community-acquired pneumonia patients with machine learning: A retrospective analysis of routinely collected health data. Value in Health. 2019 Nov;22:S882.

[43] Wu C, Rosenfeld R, Clermont G. Using Data-Driven Rules to Predict Mortality in Severe Community Acquired Pneumonia. PLoS ONE. 2014;9(4).

[44] Kim J, Choo H, Shin SY, Song KD. Synthesis and quality assessment of combined time-series and static medical data using a real-world time-series generative adversarial network. Sci Rep. 2024 Aug 17;14(1):19064. doi: 10.1038/s41598-024-69812-7. PMID: 39154144; PMCID: PMC11330441.

[45] Suter-Widmer, I., Christ-Crain, M., Zimmerli, W. et al. Predictors for length of hospital stay in patients with community-acquired Pneumonia: Results from a Swiss Multicenter study. BMC Pulm Med 12, 21 (2012). 10.1186/1471-2466-12-21

