## Supplementary material for "Predicting community-acquired pneumonia outcome using time series data and machine learning": ICD-10 codes included in the research

Supplementary File A:

Extracted from [20]

**ICD10 codes used to extract CAP episodes**

A01.0, A02.2, A20.2, A21.2, A22.1, A31.0, A37.0, A37.1, A43.0, A48.1, B01.2, B05.2, B37. 1, B38.0, B38.1, B38.2, B39.0, B39.1, B39.2, B58.3, B59, B77.8, J12.0, J12.1, J12.2, J12.3, J 12.8, J12.8, J12.9, J13, J14, J15.0, J15.1, J15.2, J15.3, J15.4, J15.5, J15.6, J15.7, J15.8, J15.9, J16.0, J16.8, J17.0, J17.1, J17.2, J17.3, J17.8, J18.0, J18.1, J18.8, J18.9, J85.1, J85.2, B59.
