## Supplementary material for "Predicting community-acquired pneumonia outcome using time series data and machine learning": Ranges of clinical utility used for the study

Supplementary File B:

**Data Cleaning and filtering ranges**

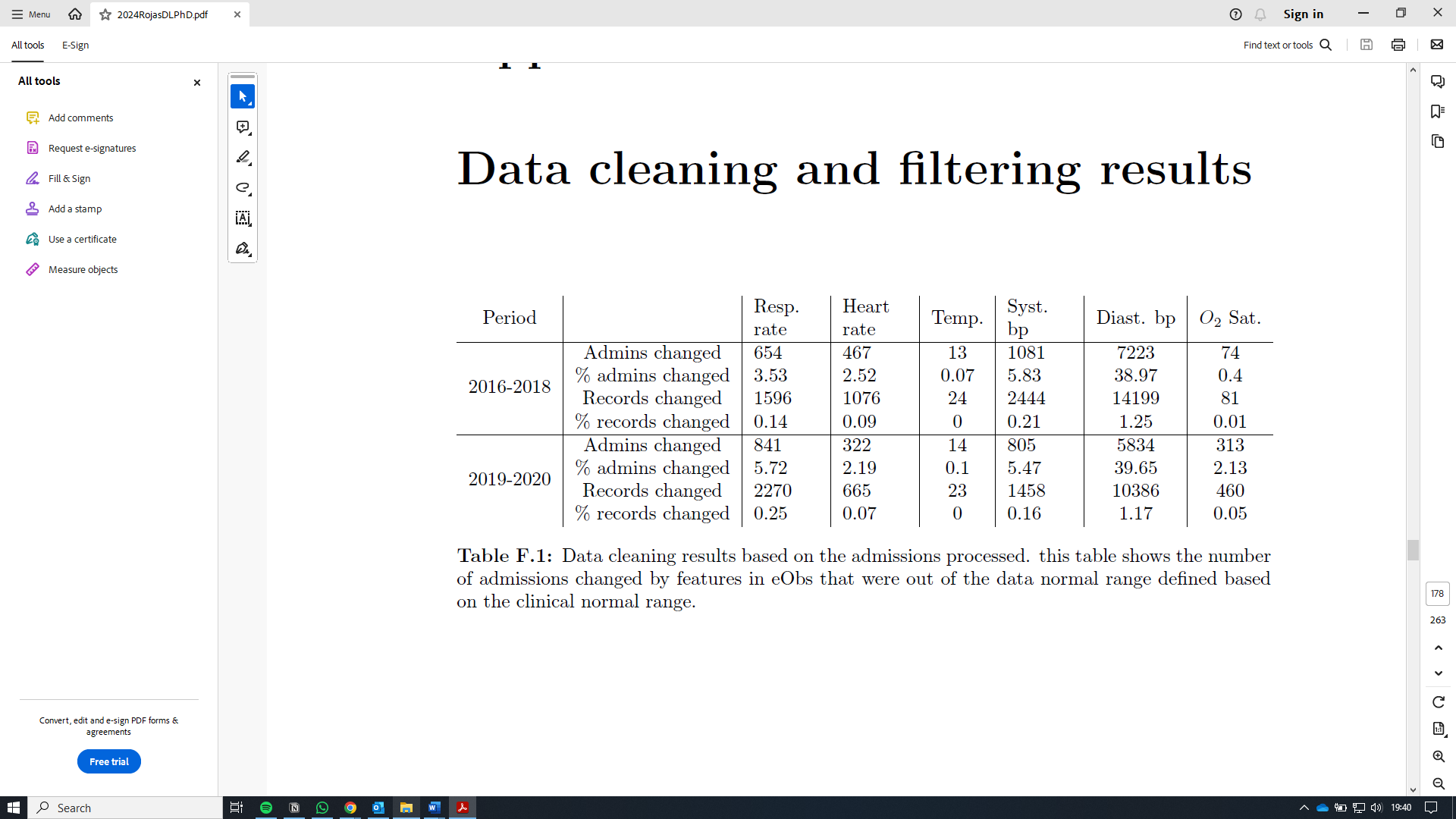

Data cleaning results based on the admissions processed. this table shows the number

of admissions changed by features in vital signs that were out of the data normal range defined based

on the clinical normal range.

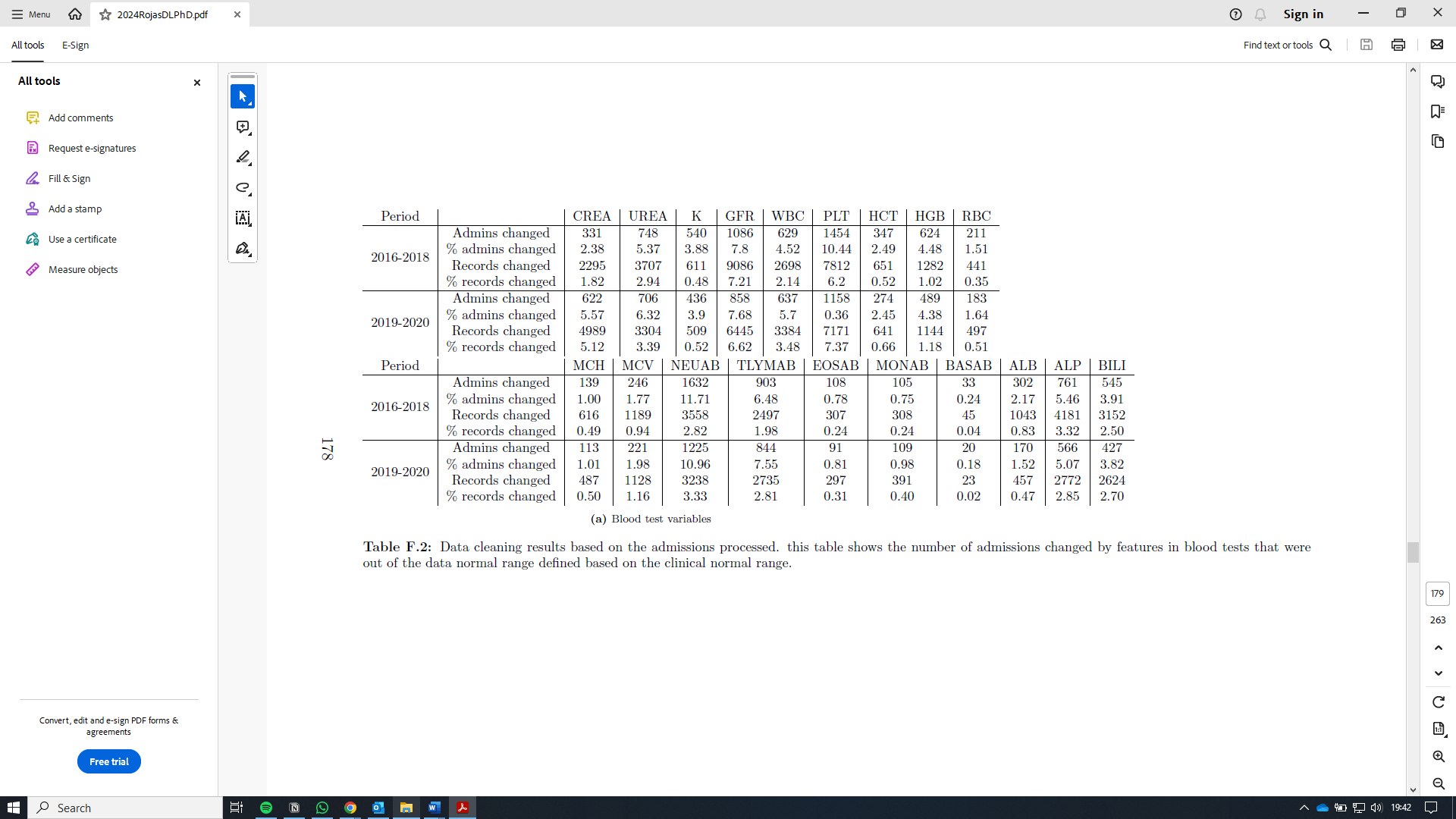

Data cleaning results based on the admissions processed. this table shows the number of admissions changed by features in blood tests that were

out of the data normal range defined based on the clinical normal range
