## Supplementary material for "Predicting community-acquired pneumonia outcome using time series data and machine learning": GitHub link for scripts

Supplementary File C:

**Data repository Link**

CAP - AI GitHub Repository. All scripts and graphs are presented in the following

link. No data are available. The study does not have ethical approval that

allows sharing the data used to produce the conclusions in this paper.

https://github.com/dflozano1172/PhD/tree/master/CAP AI Repository
