## Supplementary material for "Predicting community-acquired pneumonia outcome using time series data and machine learning": Results by characteristics age stratification

Supplementary File D:

**Data repository Link**

Age stratification by classifier and collection

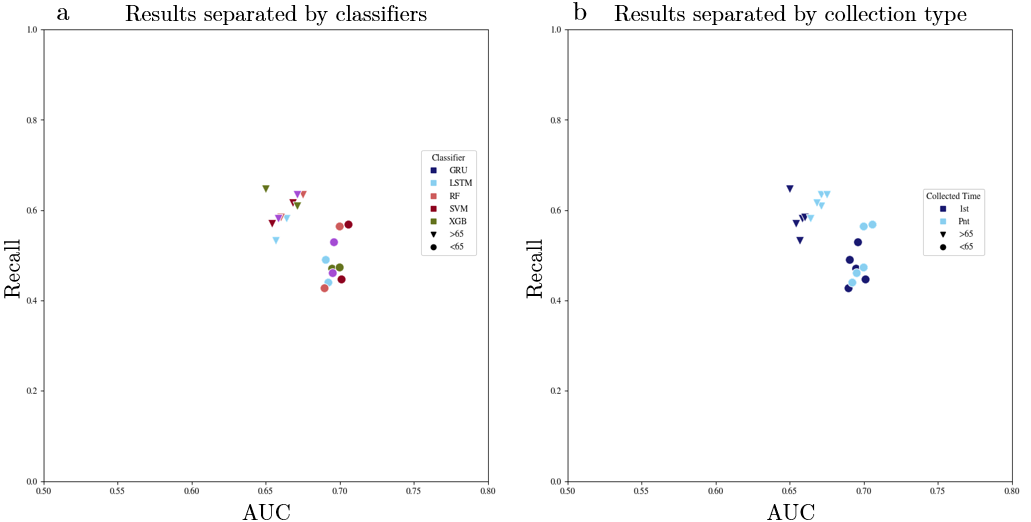

Results for Age stratification. (a) all the models created for this age stratification balanced dataset produced evident better results that unbalanced. (b) results divided by classifier aspect of models that used balanced datasets, and (c) results divided by collection time aspect of models that used balanced datasets.
