## Supplementary material for "Predicting community-acquired pneumonia outcome using time series data and machine learning": hyper-parameters and values of models implemented

### Hyperparameters considered in the fine-tuning process while training.

These are the python codes used to determined the hyper-parameters to fine tune during the training process.

#### 1. SVM model

model = svm.SVC(probability = True, random_state = 0, max_iter = 1000000)

kernels = ['linear', 'poly', 'rbf', 'sigmoid']

\# define grid ------------------------

weights = [{1:1}, {1:2}, {1:3}, {1:4},{1:5},{1:8},{1:10}, {1:25},{1:50},{1:75},{1:99},{1:100},{1:500}]

param_grid = dict(class_weight =weights, kernel =kernels)

#### 2. Random Forest

model = RandomForestClassifier(max_features = None, n_jobs=-1, bootstrap = True, random_state = 0)

\# define grid ------------------------

weights	= [{1:1}, {1:2}, {1:3}, {1:4},{1:5},{1:8},{1:10}, {1:25},{1:50},{1:75},{1:99},{1:100},{1:500}]

criteria	= ['gini', 'entropy']

depth	= [10, 20, None]

param_grid = dict(class_weight =weights, criterion = criteria, max_depth = depth)

#### 3. Extreme Gradient Boost

model = XGBClassifier(use_label_encoder=False, objective = 'binary:logistic', eval_metric = 'auc')

\# define grid ------------------------

weights	= [1, 2,3,4,5, 8, 10, 25, 50, 75, 99, 100, 500]

boosters	= ['gbtree', 'gblinear']

lambdas	= [1, 5,10, 100]

alphas	= [0, 10, 100]

param_grid = dict(scale_pos_weight=weights, booster = boosters, reg_lambda = lambdas, reg_alpha = alphas)

#### 4. Long short-term memory

clf == 'LSTM':

batch_size	= [10, 20, 60, 100, 250, 500]

epochs	= [10, 50, 100, 500]

dropout_rate = [0.2, 0.5]

param_grid = dict(batch_size=batch_size, epochs=epochs, dropout_rate=dropout_rate)

X_train = X_train.reshape(len(X_train),1,X_train.shape[1]); X_valid = X_valid.reshape(len(X_valid),1,X_valid.shape[1])

def create_model(dropout_rate = 0.0):

model = Sequential()

model.add(layers.LSTM(80, activation='relu', input_shape=(1, X_train.shape[2])))

model.add(layers.Dropout(dropout_rate))

model.add(layers.Dense(10, activation='relu'))

model.add(layers.Dropout(dropout_rate))

model.add(layers.Dense(1, activation='sigmoid'))

model.compile(loss='binary_crossentropy', optimizer='adam')#, metrics=['accuracy'])

return model

model = KerasClassifier(build_fn=create_model, verbose=0)

#### 5. Gated recurrent Unit

elif clf == 'GRU':

batch_size	= [10, 20, 60, 100, 250, 500]

epochs	= [10, 50, 100, 500]

dropout_rate = [0.2, 0.5]

param_grid = dict(batch_size=batch_size, epochs=epochs, dropout_rate=dropout_rate)

X_train = X_train.reshape(len(X_train),1,X_train.shape[1]); X_valid = X_valid.reshape(len(X_valid),1,X_valid.shape[1])

def create_model(dropout_rate = 0.0):

model = Sequential()

model.add(layers.GRU(80, activation='relu', input_shape=(1, X_train.shape[2])))

model.add(layers.Dropout(dropout_rate))

model.add(layers.Dense(10, activation='relu'))

model.add(layers.Dropout(dropout_rate))

model.add(layers.Dense(1, activation='sigmoid'))

model.compile(loss='binary_crossentropy', optimizer='adam')#, metrics=['accuracy'])

return model
